# Feedback Processing as it Relates to Suicidal Ideation and Behavior Using a Time-Frequency Approach

**DOI:** 10.1101/2023.12.06.23299613

**Authors:** Asha Pavuluri, Jessica Ellis, Anthony Vivino, Devin Butler, Melanie Arenson, Thomas Joiner, Brad Schmidt, Edward Bernat

**Affiliations:** University of Maryland, College Park; Technical Resources International, LLC; US Department of Veterans Affairs; Boston University; Florida State University

**Keywords:** suicidal ideation, suicidal behavior, feedback processing, time-frequency analysis

## Abstract

**Background:** Suicidal ideation (SI) and suicidal behavior (SB) may have different etiological pathways, particularly related to feedback processing. Identifying ERP components related to these suicide states may help researchers localize and understand these differences. ***Objective.*** Our primary purpose was to utilize time-frequency decomposition techniques to assess neurophysiological correlates of SI and SB during positive and negative feedback processing.

**Methods:** 271 subjects (55.7% female; mean age=35.75, SD=16.07; 30.6% veterans) completed a questionnaire battery and a multi-task protocol, including a gambling feedback task (Gehring & Willoughby, 2002), while electroencephalography (EEG) was collected using a 96-channel EEG system. Exploratory factor analysis (EFA) was conducted across 20 self-report suicide items selected to index SI and SB from several suicide-relevant questionnaires. EFA results produced 2 factors, corresponding to SI and SB. Time-frequency principal component analyses produced measures across all frequency bands (i.e., delta-FN/P3, delta-SW, theta-FN, alpha, beta-1, beta-2, and gamma).

**Results:** Excepting alpha amplitude, all measured frequencies were related to SB, and not SI (across loss and gain, with some loss-gain differences). Alpha, uniquely, showed a relationship to SI and not to SB (across loss and gain, no loss-gain differences). Robust regressions confirmed that the delta, theta, and high-frequency (HF; beta-1, beta-2, and gamma as combined HF loss, gain, and loss-gain difference factors) measures were each independently related to SB.

**Discussion:** Broadly, the results indicate that individuals with SB showed heightened neurophysiological response across multiple ERP components (except alpha) to gambling feedback, with significantly greater increases to loss stimuli relative to gain stimuli.

**General Scientific Summary:** This study, using time-frequency EEG/ERP measures from a gambling feedback task, supports the theoretical framework that suicidal behavior and suicidal ideation can be indexed separately. Generally, stronger reactivity to all feedback (i.e., loss and gain feedback during a gambling task) is seen in those with suicidal behavior and not in those with suicidal ideation, but with a small separate effect related to suicidal ideation.

## Introduction

Every year, more than 700,000 people die by suicide worldwide (World Health Organization, 2021), and it is the tenth leading cause of death overall (Centers for Disease Control and Prevention (CDC), 2022; National Center for Injury Prevention and Control, 2019). Suicidal ideation (SI) refers to thinking about, considering, or planning suicide assessed in terms of frequency, intensity, and quality of such thoughts (CDC, 2022; Klonsky & May, 2010). Suicidal behaviors (SB) refer to the actual intent associated with such thoughts, with an intent to die, that can potentially be dangerous and lethal (CDC, 2022; Klonsky & May, 2010). Despite billions spent in research funding, fairly little is known about what differentiates individuals who have SI from those who then engage in SBs (Kessler et al., 2005; Klonsky & May, 2014; May & Klonsky, 2016; Nock et al., 2008). Thus, there is strong motivation to better understand parameters differentiating patients who present with SI versus those that subsequently present with SB, and to identify relevant mechanisms that will allow clinicians to prioritize and appropriately intervene when a person is contemplating suicide.

### Differentiating SI and SB

It is well understood that SI and SB have different etiological pathways (Klonsky & May, 2010). To elaborate, there are two related issues in the field to address: 1) a strong understanding of SI, but not SB, and 2) a weaker understanding of what differentiates SI from SB and the progression from SI to SB.

The current understanding of SB suggests that capability for suicide, from the Interpersonal Theory of Suicide (Joiner 2005; cf. Klonsky et al., 2018), and deliberate self-harm (Duarte et al., 2020) are well-established attributes that are more specific to attempters. Previous studies found diagnoses and traits such as depression, hopelessness, most mental disorders, impulsivity, anhedonia, low belongingness, or burdensomeness (all established markers of “suicide”) are robust predictors of SI, but not SB (Klonsky & May, 2014; Kessler et al., 1999; May & Klonsky, 2016; Klonsky et al., 2017; Klonsky et al., 2018). A valuable addition to suicide theory would be establishing indicators unique to SB.

While it is known that previous suicidal thoughts are significant predictors of later thoughts and behaviors, the size of this effect is less established (Ribeiro et al., 2016); over 90% of those reporting SI do not progress to SB within the next two years (ten Have et al., 2009). A comprehensive 50-year meta-analysis of 365 studies of suicidal thoughts and behaviors risk factors found that, currently, prediction based on known risk factors remains at above-chance but sub-optimal levels (Franklin et al., 2017). Furthermore, recent research suggests most known psychosocial markers of “suicide” are in fact markers of SI and do little to identify progression to SB (Acosta et al., 2012; Klonsky & May, 2010, 2014; Naifeh et al., 2019). The importance of successfully predicting those at highest risk of moving from SI to SB remains a largely unanswered question (Klonsky & May, 2014).

Overall, several studies have noted the need for more dependable differentiation of SI and SB through existing questionnaires, characteristics, or mechanisms (Baik et al., 2018; Klonsky et al., 2018). Addressing this differentiation can provide clinicians with more clear and dependable delineations between SI and SB.

### Suicide-Related Neurophysiological Mechanisms

Recently, researchers have investigated processing deficits to better understand suicidal frameworks. Paired with increased interest in neurobiological markers of mental health difficulties, investigators have employed fMRI and EEG techniques to study decision-making processes, as impairments in reward and feedback processing have been implicated in suicidal thoughts and behaviors, as well as in the lethality of attempts (for review, see Dombrovski & Hallquist, 2017; Bani-Fatemi et al., 2018; Jollant et al., 2011; Van Heeringen et al., 2011; de Aguiar Neto & Rosa, 2019; Gallyer et al., 2021).

fMRI studies have revealed that those with SI and/or SB display increased anterior cingulate cortex (ACC) activation and reduced orbitofrontal and dorsolateral activity (Van Heeringen et al., 2011). Furthermore, those with prior SB displayed reduced functional connectivity between the ACC and prefrontal regions (for review, see Bani-Fatemi et al., 2018; Jollant et al., 2011). Dombrovski and colleagues (2013) found that older adults with a history of SB had a blunted response in paralimbic structures and the ventromedial prefrontal cortex to high expected rewards relative to those without prior SB, suggesting patients with prior SB experience disruptions in reward processing. These results suggest that deficits in performance monitoring and feedback processing, broadly, may relate to understanding the neural associations of SI and SB.

Event-related potentials (ERP, i.e. electrocortical data) provide high temporal resolution data that are cost-effective to collect relative to fMRI studies. ERP research related to SI and SB has revealed: lower serotonergic activity in participants with SB compared to no SB (Kim & Park, 2013; Chen et al., 2005); reduced Late Positive Potential (LPP) in SB to threat images compared to no suicide history and SI (Albanese et al. 2019; Weinberg et al., 2017); and, of interest to this study, feedback processing related to psychological pain and risk factors in suicidality, indicated by P200, P300 (P3), and Reward Positivity measures (RewP) (Song et al., 2019; Tsypes et al., 2021).

Few studies have used EEG to investigate feedback processes related to SI and/or SB. Song et al. (2019) compared feedback processing between those with major depressive disorder and low suicide risk (MDD-LSR) or high suicide risk (MDD-HSR) to healthy controls and found that the feedback-P3 to positive and negative feedback in healthy controls was greater relative to MDD-LSR or MDD-HSR groups, with no difference between the MDD groups. Tsypes et al., found a blunted cue-P3 in a monetary incentive delay task for suicide attempters versus non-attempters (2021). Still, a meta-analysis of 27 studies found that the current literature is severely underpowered in terms of sample size (with Ns of 10-100), with mixed results for ERP and SI and SB outcome combinations (Gallyer et al., 2021). Even fewer studies have examined high-frequency (HF) measures, such as alpha, beta-1, beta-2, and gamma related to suicide and/or feedback processing. One study measured an increase in alpha spectral power in the parieto-occipital area for those with suicidal attempts compared to those with non-suicidal self-injuries (Iznak et al., 2021). In another, non-suicidal depressed patients displayed decreased beta and low gamma activity in the frontal regions (Benschop et al., 2019). During feedback processing, others observed a large increase of beta-gamma oscillatory activity only after unexpected monetary gains, irrespective of the magnitude of the reward (HajiHosseini et al., 2012). Despite these relevant studies, HF oscillatory activity elicited during feedback processing is less understood. By utilizing EEG and time-frequency (TF) measures, a more comprehensive understanding of neural mechanisms related to suicide can be developed.

### Time-Frequency Analysis

The current project utilizes a TF approach: this involves a unique parsing apart of different frequencies from the time-domain data. The feedback negativity (FN) ERP component is a well-established physiological marker of outcome evaluation found in the medial-frontal region that peaks approximately 250 milliseconds (ms) after feedback and differentiates negative and positive feedback (i.e., RewP) (Gehring & Willoughby, 2002). For instance, while it was traditionally posited as a relatively low-level response to negative outcomes, recent research using TF analysis has revealed that the FN is comprised of two overlapping parts: (1) a negative-going, loss-sensitive theta signal, and (2) a positive-going, gain-sensitive delta signal (Bernat et al., 2011; Bernat et al., 2015; Yeung, 2004). The ability to separate these pieces has been critical in understanding how different aspects of feedback processing relate to psychopathology such as anxiety (Ellis et al., 2018) and externalizing disorders (Bernat et al., 2011). Importantly, while the FN is the primary component typically assessed in feedback paradigms, recent research has shown that early attention orienting and later low wave activity maybe implicated as well (Ellis et al., 2018). Few studies have used TF measures in the context of suicide and adding this perspective may help identify relevant neural mechanisms.

### Current Study

The current study aimed to increase existing understanding of feedback-relevant ERP measures related to suicide with a comprehensive analysis of TF amplitude measures in a gambling feedback task. Given previous theoretical understandings of SI and SB, we hypothesized that those presenting with SB will have a stronger response to both loss and gain feedback than those presenting with SI (Klonsky & May, 2014).

## Methods

### Participants

281 participants were recruited from a Southeastern American University. Eight participants were excluded during Data Preprocessing due to incomplete data (five participants missing data files) or unusable data (three participants removed because >25% of trials rejected due to unreadable EEG artifacts). Two additional participants were excluded due to missing suicide factor loadings (i.e., missing questionnaire data), leaving 271 participants for data analysis (55.7% female; mean age=35.75, SD=16.07; 30.6% veterans; 59.4% Caucasian, 26.6% Black, 2.6% Asian, 0.4% Pacific Islander, 0.4% American Indian/Native American, 10.7% Other; 9.6% Hispanic; 53.9% Some college/two-year degree).

### Procedure

#### Screening Appointment

Participants who were deemed potentially eligible, based on an initial phone screen, were invited to complete a more comprehensive screening appointment. During this appointment, participants underwent a diagnostic interview (First et al., 2015) with a trained therapist, and a thorough suicide risk assessment (Joiner et al., 1999). After the diagnostic interview, participants completed self-report measures to help determine study eligibility and inform diagnostic decisions. If participants were deemed ineligible based on the screening appointment they were thanked for their time and given relevant community referrals based on their needs.

#### Baseline Appointment

Participants completed a battery of self-report questionnaires. Upon completion, they were scheduled for a baseline neurophysiology assessment and provided with monetary compensation. Only baseline data was used for the purposes of this study.

#### Questionnaires

Participants were asked to complete a battery of demographic and psychological questionnaires. The following questionnaires contained items that evaluate either SI or SB, or both:

- *Beck Scale for Suicide Ideation (BSS),* a 21-item scale that assesses an individual’s current suicidal attitudes and behaviors (Beck et al., 1979).

`o` Internal consistency coefficient (𝛼=.89).
`o` Interrater reliability *Κ*=.83 (*p*<.001).
`o` Construct validity with hopelessness and depression (*r*=.47, *p*<.001 and *r*-.39, *p*<.001, respectively), when Beck Depression Inventory was parsed out still *r*=.32, *p*<.001).
- *Suicide Behaviors Questionnaire-Revised (SBQ-R),* a 4-item questionnaire that aims to evaluate suicide risk behaviors (Osman et al., 2001).

`o` For psychiatric adult inpatient sample, intercorrelations ranged from *r*=.62 (likelihood to threat) to *r*=.76 (past attempts vs. frequency), with 𝛼=.87.
`o` Four item questionnaire developed from original 30-item SBQ (Linehan, 1981).
- *Depressive Symptom Inventory-Suicidality Subscale (DSI-SS),* a 4-item suicide scale that is widely used to detect and prevent suicide (Metalsky & Joiner, 1997).

`o` Internal consistency measured 𝛼=.90.
`o` Strong construct validity seen through correlations between suicidality and depressive symptoms (*r*=.60, *p*<.0001), and between suicidality and general distress (*r*=.49, *p*<.0001) (Joiner et al., 2002).
`o` Developed as subscale from Hopelessness Depression Symptom Questionnaire, 32-item self-report measure (Abramson et al., 1989).
- *Suicide History Form (SHF),* a series of 6 questions to understand amount, extent, and recency of previous suicidal attempts.
- *Beck Depressive Inventory-II (BDI-II),* a 21-item questionnaire to score seriousness of depressive symptoms (item related to suicide selected from questionnaire) (Beck et al., 1996).

`o` The test-retest reliability of the BDI-II ranged from *r*=.73 to *r*=.92 (Wang & Gorenstein, 2013).
`o` The internal consistency of the BDI-II was 𝛼=.90.
- *Suicide Intent Scale (SIS),* a 5-item questionnaire meant to understand intention to attempt suicide and extent of previous suicide attempts (Beck, Schuyler, & Herman, 1974).

`o` Interrater reliability for 45 attempted suicides was *Κ*=.95 (Beck, Morris, & Beck, 1974).

#### Gambling Task

The gambling task was a modified version of the Gehring and Willoughby (2002) task, in which participants would be presented with two monetary values on the screen and be given feedback about whether they won or lost money after selecting one of the values (Figure 1). The four different combinations of numbers could be of equal or different cent values (i.e., 5 and 5, 5 and 25, 25 and 5, 25 and 25) with a random pattern of winning or losing (to prevent the possibility of prediction). Feedback was presented for 1000 ms after selection by the chosen box turning either red or green (with the color indicating winning being counterbalanced across participants) and the unselected box would turn, either, the other color or the same color between red and green (i.e., it is possible that both values could have been losing selections or winning selections). Participants completed 7 blocks (each block contained two sets of these 16 trial types, randomly ordered) of 32 trials each, for a total of 224 trials. After each block was completed, participants received feedback regarding their win/loss ratio for that particular block.

**Figure 1.**
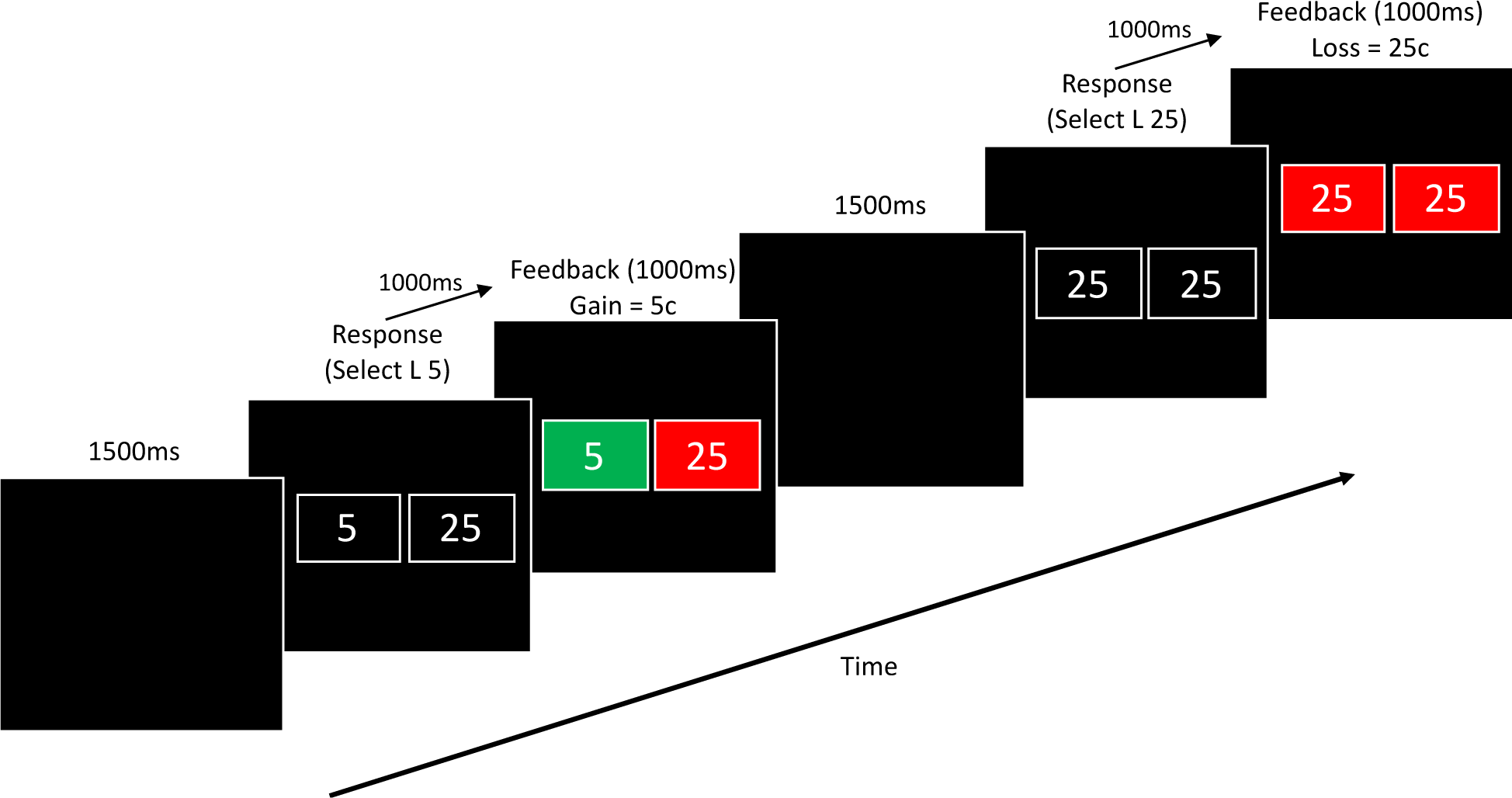
Sequence of Stimuli and Feedback Events in the Gambling Task.

### Exploratory Factor Analysis

Exploratory factor analysis (EFA) was used to interpret SI and SB factors. An iterative EFA process was conducted in SPSS (V 29.0.0.0, IBM Corp.) across suicide related items from the questionnaires previously listed. 20 self-report suicide items were selected to index SI and SB. Principal Axis Factoring (PAF) was used with an orthogonal varimax rotation, which would maximize independence of loadings and factors. This was important to account for any overlapping traits between SI and SB, and to develop a factor matrix with a more uncorrelated structure.

### Psychophysiological Data Collection

Neurophysiological data was recorded using a BrainVision 96-channel actiCap (sintered Ag-Ag/Cl; international 10-20 system; Jasper 1958) as well as a 24-bit battery-supplied compatible amplifier (Neuroscan Inc.). Horizontal electrooculogram activity was recorded from

electrodes placed on the outer canthus of both eyes, while vertical electrooculogram activity was recorded from electrodes placed above and below the left eye (note: four of the 96 EEG channels are repurposed for this rather than using other peripheral electrodes). Impedances were kept below 10 kΩ. EEG signals were vertex referenced (FCz) during recording. Recordings were collected using a 500 Hz sampling rate, analog 0.05 to 100 Hz bandpass filter.

### Data Processing

#### Preprocessing

EEG data preprocessing was conducted using a custom MATLAB script (version 9.3.0.713579 [R2017b]; The MathWorks, Inc.) utilizing both original and EEGLAB functions (Garland et al., 2021). ERP analysis was performed in the Psychophysiology Toolbox (PTB) developed by Dr. Edward Bernat (Buzzell et al., 2022).

The preprocessing steps are set up to clean and correct the EEG datafiles. The output set of preprocessed mat files that can then be analyzed through the PTB. Epochs of 3,000 ms were taken from 1,000 ms pre to 2,000 ms post-stimulus with a 500 ms pre-stimulus baseline correction and 100 ms post-stimulus baseline correction. Electrodes were re-referenced to averaged mastoid sites. Data were corrected for ocular artifacts (Gratton et al., 1983) and downsampled to 256 Hz using the MATLAB resample function (Mathworks, Inc.), which applied an anti-aliasing filter during resampling. The respective triggers for each task were set in the respective task preprocessing script (i.e., loss and gain trials in the gambling task).

A low-pass filter of 50 Hz was applied to the continuous data, and then ERP epochs were created as described previously. Epochs were computed separately for loss and gain trials. For each individual, epochs were ranked according to number of extreme (>±150 mV) data points across all channels, and the worst 5% of epochs were removed. In addition, individual channels were interpolated across all data if they exceeded the threshold of 5 SDs in the domains of kurtosis and activity probability. After baseline (500–100 ms before stimulus) correction, each epoch was evaluated separately and channels with extreme (>±150 mV) data points were interpolated only for that epoch, while epochs with more than two bad channels were rejected and removed from the data. A final visual inspection was conducted to remove epochs with unusual artifacts. ERP component scores were extracted and exported for further statistical analyses.

### Data Reduction

#### Time-Frequency Analysis

Using the PTB, developed by Dr. Edward Bernat, principal components analysis (PCA) was conducted on frequency filtered TF surfaces of the EEG/ERP data (see Buzzell et al, 2022 for a detailed treatment of this approach). The data analyzed are those time-locked to the gain and loss feedback, 100 ms after a selection is made. Data were analyzed separately for SI and SB factors, by loss and gain feedback ERPs, and loss-gain differences.

#### Time-Frequency Components: Delta, Theta, Alpha, Beta-1, Beta-2, and Gamma

TF-PCA was used to identify distinct frequency activity (for details, see Bernat et al., 2005; Bernat et al., 2011; Nelson et al., 2011; Bernat et al., 2015; Buzzell et al., 2022; and Watts et al., 2018). Figure 2 provides the grand-average TF-PCA decomposition for the six frequency bands with there being two components extracted for delta, and one component extracted for the remaining frequency bands. Scree plots identified the ideal components to review for each frequency band.

**Figure 2.**
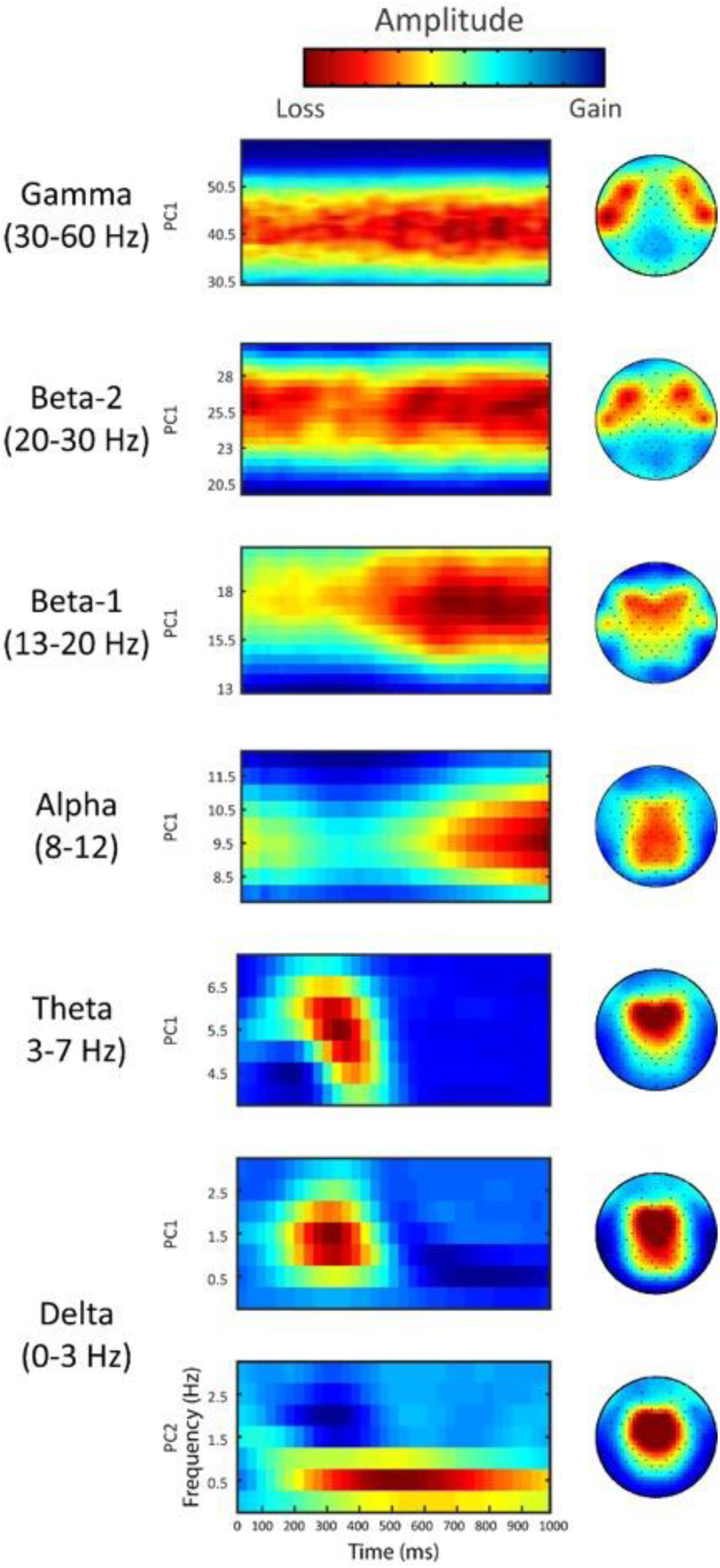
Delta PC1 and PC2, Theta, Alpha, Beta-1, Beta-2, and Gamma Grand Averages.

**Figure 3.**
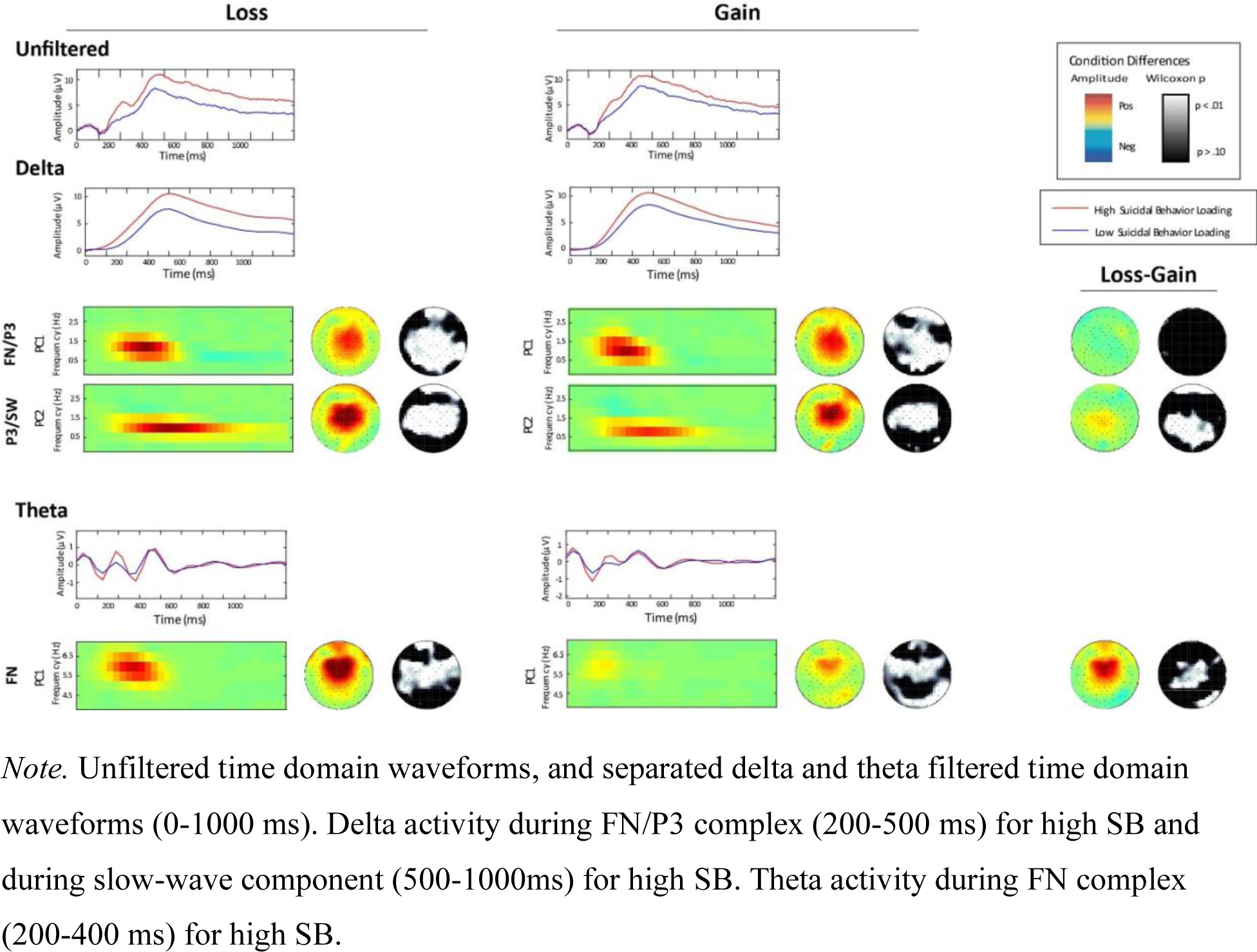
Delta and Theta Time-Frequency Analysis Related to Suicidal Behavior.

In low-frequency (LF) bands, two PCs were identified for delta and one PC was identified for theta. The first delta (0-3 Hz) component occurred during the FN-P3 complex (200-500 ms post-feedback) and the second represented the slow-wave (SW; 500-1000 ms), and the theta (3-7 Hz) component was consistent with the timing of the FN time window (200-400 ms). Given previous relevant work (Watts et al., 2018) and PCs extracted in this study, the delta center of activation was evaluated as a CZ-CPZ combination. The delta FN-P3 measure accounted for 62.59% of the variance, with the delta-SW measure accounting for 9.84% of the variance in the gambling data. For similar reasons as stated for delta, theta activation was chosen to be evaluated as a FCZ-CZ combination. The theta measure accounted for 54.03% of the variance.

In HF bands, a single component for each measure of alpha (8-12 Hz), beta-1 (13-20 Hz), beta-2 (20-30 Hz), and gamma (30-60 Hz) were identified. For alpha, the activity appears to begin approximately 500 ms post-feedback. Given previous work (Iznak et al., 2021) and current findings, alpha activation was evaluated at POZ. The alpha PCA accounted for 64.89% of the variance. Although beta-1 activity, like alpha activity, begins closer to 500 ms post-stimulus, beta-2 and gamma activity began earlier with both appearing to be continuous and consistent measure from feedback onset. Beta-1, beta-2, and gamma activity was evaluated as a symmetrical, bilateral, fronto-temporal electrode cluster of F7-FT7-F8-FT8 (Satapathy et al., 2019). Given persisting activity from −1000 to 2000 ms for beta-1, beta-2, and gamma, a robust regression was conducted in 500 ms blocks, but no significant unique effect was found among these blocks; therefore, 0-1000 ms windows were used in analysis. The beta-1, beta-2, and gamma measures, respectively, accounted for 65.84%, 75.80%, and 67.74% of the variance.

PCA was then applied across the full set of TF condition averages to a post-stimulus time window of 0–1000 ms, following methods previously mentioned (Buzzell et al., 2022).

#### Gambling Results without Suicide Conditions

The grand average TF decompositions across frequencies presented in Figure 2 depict the basic gambling effects.

### Data Analysis

There were two main parts to the data analyses: 1) identifying the relationships between suicide factors and TF activity across the frequencies, and 2) understanding the shared and unique variance among the observed effects. First, Spearman correlations were calculated between each of the suicide factors and delta, theta, alpha, beta-1, beta-2, and gamma frequencies in SPSS. Second, robust regressions (conducted in Stata/SE 17.0 using *rreg* command) were fit to examine the unique versus shared variances in the frequency effects associated with the SB factor.

#### EFA: High-Frequency Latent Factor

Given the similarity in the location and activity among the HFs (beta-1, beta-2, and gamma), EFAs were conducted for loss, gain, and loss-gain differences to analyze the relationship among these frequencies. The loss factor explained 91.78% of the variance in the data, with beta-1, beta-2, and gamma having respective loadings of .91, .96, and .95. The gain factor explained 91.11% of the variance in the data, with beta-1, beta-2, and gamma having respective loadings of .88, .96, and .95. Finally, the loss-gain factor explained 60.25% of the variance in the data, with beta-1, beta-2, and gamma having respective loadings of .45, .83, and .64. The HF Factors (HF loss factor, HF gain factor, and HF loss-gain differences factor) were used in the robust regression analyses.

## Results

### Exploratory Factor Analysis

EFA using PAF with a varimax rotation produced 2 latent factors (explaining a total of 56.06% of the variance), corresponding to SI (15 items, 45.20% of variance) and SB (5 items, 12.86% of variance), respectively (Table 1).

**Table 1.**
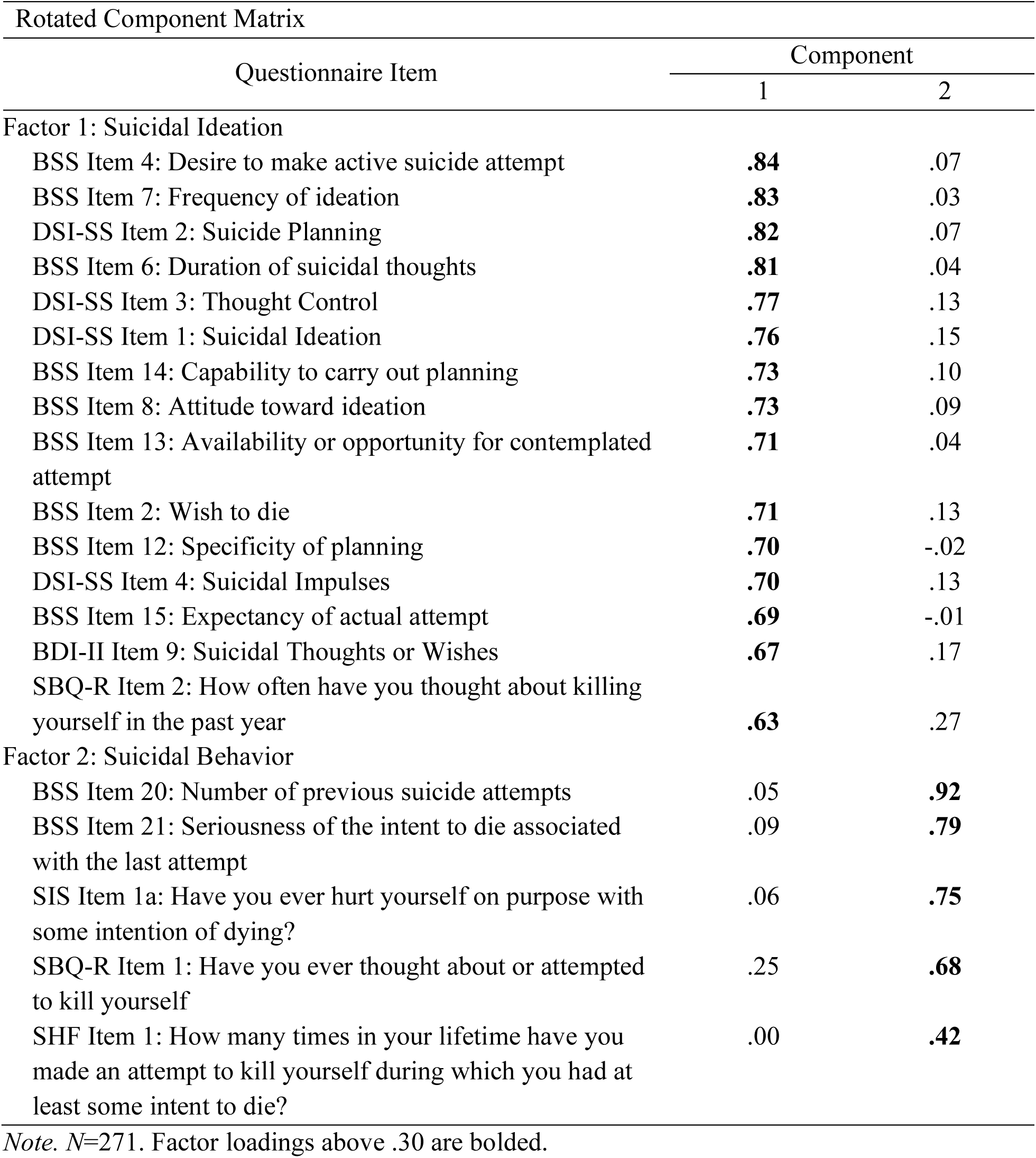
Exploratory Factor Analysis Conducted Across 20 Items to Index Suicidal Ideation and Suicidal Behavior.

Questionnaire items related to SI or SB were selected and an iterative process of removing items that either did not load highly onto either factor (i.e., loading less than .30) or did not load clearly onto one factor over the other (i.e., similarly loaded highly or poorly onto both factors) was conducted. This left the 20 items listed in Table 1 to develop the SI factor (component 1) and the SB factor (component 2).

### Psychophysiology Results

#### Delta and Theta

Delta evoked power was centro-parietal (average electrodes CZ-CPZ) and theta evoked power was medial-frontal (average electrodes FCZ-CZ). Delta-FN/P3 complex displayed significant differences between high and low SB during loss (*r*=.20, *p*<.001) and gain (*r*=.16, *p*<.01) trials, but no significant loss-gain differences (Table 2). Delta-SW component displayed significant differences between high and low SB during loss (*r*=.26, *p*<.01) and gain (*r*=.18, *p*<.01) trials, as well as loss-gain differences, favoring the loss trials (*r*=.18, *p*<.01) (Table 2). Theta-FN component displayed significant differences between high and low SB during loss (*r*=.16, *p*<.01) and gain (*r*=.13, *p*<.05) trials, with loss-gain differences, favoring loss trials (*r*=.15, *p*<.05) (Table 2).

**Table 2.** Delta (CZ-CPZ) and Theta (FCZ-CZ) Spearman Correlations for Loss and Gain Feedback by ERP Component.

| Condition | Ideation |  |  | Behavior |  |  |
| --- | --- | --- | --- | --- | --- | --- |
|  | Delta-FN/P3 | Delta-SW | Theta-FN | Delta-FN/P3 | Delta-P3/SW | Theta-FN |
| Loss | .05 | .02 | .07 | **.20\*\*\*** | **.26\*\*\*** | **.16\*\*** |
| Gain | .06 | -.02 | .08 | **.16\*\*** | **.18\*\*** | **.13\*** |
| Loss-Gain | -.03 | .05 | .02 | -.02 | **.18\*\*** | **.15\*** |

† .10 ≤ p < .05, \*p ≤ 0.05, \*\*p ≤ 0.01, \*\*\*p ≤ 0.001

#### Alpha

Total power PCA revealed a significant relationship between SI and feedback processing for alpha frequency band, but not for SB (Figure 4). Parieto-occipital effects (POZ) from 0-1000 ms were identified in loss (*r*=.16, *p*<.01 and gain (*r*=.15, *p*<.05) trials, but not in loss-gain differences (Table 3).

**Figure 4.**
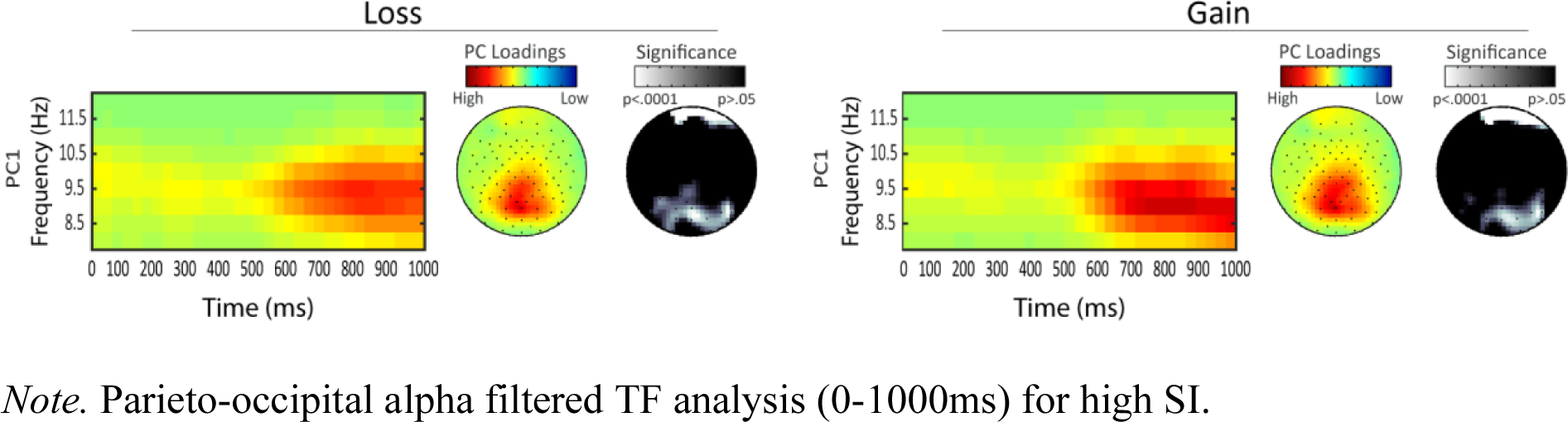
Alpha Filtered TF Analysis Related to Suicidal Ideation.

**Table 3.**
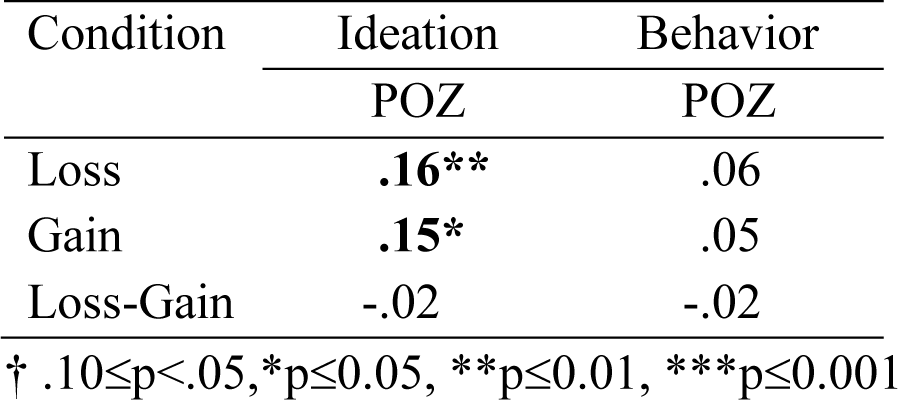
Alpha Spearman Correlations for Loss and Gain Feedback.

#### Beta 1, Beta 2, and Gamma

Total power PCA revealed a significant relationship between SB and feedback processing, but not SI and feedback processing for beta 1, beta 2, and gamma frequency bands. Bilateral, fronto-temporal (F7-FT7-F8-FT8) effects from 0-1000 ms were identified in loss and gain trials, but not in loss-gain differences, across all three frequency bands (Figure 5). Beta 1 (12-20 Hz) showed positive correlation with SB for loss (r=.25, p<.001) and gain (r=.24, p<.001) trials (Table 4). Beta 2 (20-30 Hz) showed positive correlation with SB for loss (r=.22, p<.001) and gain (r=.21, p<.001) trials (Table 4). Gamma (30-60 Hz) showed positive correlation with SB for loss (r=.22, p<.001) and gain (r=.21, p<.001) trials (Table 4).

**Figure 5.**
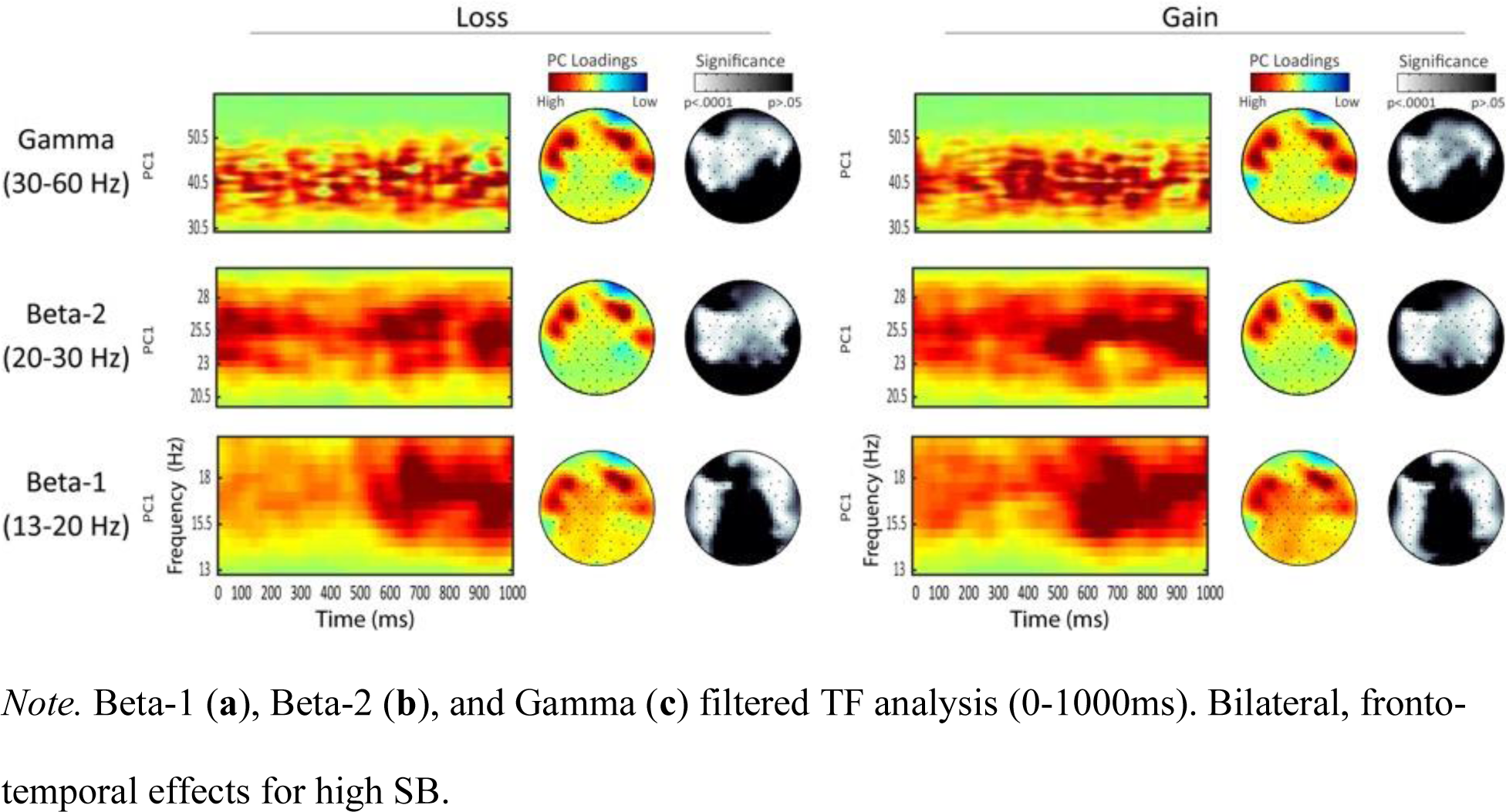
Beta-1, Beta-2, and Gamma Filtered TF Analysis Related to Suicidal Behavior.

**Table 4.** Beta-1, Beta-2, and Gamma Spearman Correlations for Loss and Gain Feedback by Suicidal Ideation and Suicidal Behavior.

| Condition | SI |  |  | SB |  |  |
| --- | --- | --- | --- | --- | --- | --- |
|  | Beta-1 | Beta-2 | Gamma | Beta-1 | Beta-2 | Gamma |
| Loss | .08 | .06 | .06 | **.25\*\*\*** | **.22\*\*\*** | **.22\*\*\*** |
| Gain | .10† | .08 | .07 | **.24\*\*\*** | **.22\*\*\*** | **.21\*\*\*** |
| Loss-Gain | -.06 | -.09 | -.08 | -.03 | -.11† | -.04 |

† .10≤p<.05, \*p≤0.05, \*\*p≤0.01, \*\*\*p≤0.001.

### Robust Regressions

Robust regressions were run in Stata to identify unique relationships with SB (note: alpha excluded because it showed relation only to SI). Of the delta components, delta-SW had the strongest impact in predicting SB (Table 5), but was still seen as unique from the other LF, theta, effects (Table 6).

**Table 5.** Suicidal Behavior and Delta Components Post-hoc Robust Regression.

| Frequency | Loss |  |  | Gain |  |  | Loss-Gain |  |  |
| --- | --- | --- | --- | --- | --- | --- | --- | --- | --- |
| | $\beta$ | $t$ | $SE$ | $\beta$ | $t$ | $SE$ | $\beta$ | $t$ | $SE$ |
| Delta-PC1 | .02 | 1.04 | .02 | .02 | 1.10 | .01 | -.03 | -.96 | .03 |
| Delta-PC2 | .03 | **2.14\*** | .02 | .03 | **1.83†** | .02 | .11 | **3.35\*\*\*** | .03 |
| Constant | -.52 | **-34.44\*\*\*** | .02 | -.52 | **-32.02\*\*\*** | .02 | -.47 | **-38.41\*\*\*** | .01 |
| F |  | **10.69\*\*\*** |  |  | **6.95\*\*\*** |  |  | **5.63\*\*** |  |

† .10≤p<.05, \*p≤0.05, \*\*p≤0.01, \*\*\*p≤0.001
Note. 95% Confidence Intervals in order of frequency measures: Loss = [-.02, .06], [.003, .06], [-.55, -.49]; Gain = [-.01, .05], [-.002, .06], [-.55, -.49]; Loss-Gain = [-.08, .03], [.05, .17], [-.50, -.45].

**Table 6.** Suicidal Behavior and Delta and Theta Post-hoc Robust Regression.

| Frequency | Loss |  |  | Gain |  |  | Loss-Gain |  |  |
| --- | --- | --- | --- | --- | --- | --- | --- | --- | --- |
| | $\beta$ | $t$ | $SE$ | $\beta$ | $t$ | $SE$ | $\beta$ | $t$ | $SE$ |
| Delta-PC2 | .04 | **4.21\*\*\*** | .01 | .04 | **3.17\*\*** | .01 | .10 | **3.42\*\*\*** | .03 |
| Theta | .06 | **1.70†** | .03 | .21 | **2.04\*** | .10 | .08 | **1.82†** | .04 |
| Constant | -.53 | **-32.87\*\*\*** | .02 | -.53 | **-31.05\*\*\*** | .02 | -.48 | **-36.52\*\*\*** | .01 |
| F |  | **11.14\*\*\*** |  |  | **8.37\*\*\*** |  |  | **6.72\*\*\*** |  |

† .10≤p<.05, \*p≤0.05, \*\*p≤0.01, \*\*\*p≤0.001
Note. 95% Confidence Intervals in order of frequency measures and constant: Loss = [.02, .06],
[-.01, .12], [-.56, -.50]; Gain = [.01, .06], [.01, .41], [-.56, -.50]; Loss-Gain [.04, .16], [-.01, .17],

An additional robust regression was conducted to identify whether a difference in the effects of LF and HF measures to SB existed based on the key measures of delta-PC2, theta, and HF factor (Table 7). The variance explained and loadings for the HF factor are described in *Data Analysis*. Although above a meaningful threshold for variance explained and EFA loadings, the lower variance percentage and loading values seen in loss-gain differences may be related to the non-significant effect seen in the HFs. Given that loss and gain factors were impactful and most relevant to the correlations in these frequencies, we proceeded with the HF Factor to represent beta-1, beta-2, and gamma.

**Table 7.** Low and High-Frequency and Suicidal Behavior Post-hoc Robust Regression.

| Frequency | Loss |  |  | Gain |  |  | Loss-Gain |  |  |
| --- | --- | --- | --- | --- | --- | --- | --- | --- | --- |
| | $\beta$ | $t$ | $SE$ | $\beta$ | $t$ | $SE$ | $\beta$ | $t$ | $SE$ |
| Delta-PC2 | .04 | **4.30\*\*\*** | .01 | .03 | **3.18\*\*** | .01 | .10 | **3.11\*\*** | .03 |
| Theta | .06 | **1.84†** | .03 | .23 | **2.24\*** | .10 | .07 | **1.63†** | .04 |
| HF Factor | .03 | **2.51\*\*** | .01 | .03 | **3.12\*\*** | .01 | -.04 | **-3.22\*\*\*** | .01 |
| Constant | -.53 | **-33.54\*\*\*** | .02 | -.53 | **-31.54\*\*\*** | .02 | -.48 | **-36.15\*\*\*** | .01 |
| F |  | **10.31\*\*\*** |  |  | **9.06\*\*\*** |  |  | **8.00\*\*\*** |  |

† .10≤p<.05, \*p≤0.05, \*\*p≤0.01, \*\*\*p≤0.001
Note. 95% Confidence Intervals in order of frequency measures and constant: Loss = [.02, .06],
[-.003, .12], [.0049, .05], [-.56, -.50]; Gain = [.01, .06], [.03, .43], [.01, .05], [-.56, -.50]; Loss-
Gain = [.04, .16], [-.0049, .17], [-.04, .01], [-.51, -.46].

## Discussion

This study aimed to better understand the differences in feedback processing associated with SI relative to SB. We found support for the following: 1) separable empirically based measures of SI and SB, and 2) distinct neural mechanisms associated with SB relative to SI.

### Differentiating SI and SB

The preliminary step was to identify a method to separately represent SI and SB. Previous research has established that there are clear etiological differences between the two (Klonsky and May, 2010), but current literature biases toward understanding SI more clearly than SB and offering limited differentiation (Klonsky & May 2014; Kessler et al., 1999; May & Klonsky 2016; Klonsky et al., 2017; Klonsky et al., 2018). Thus, we were not able to directly adopt measures from the field. The suicidality EFA conducted here helped to provide separable factors of SI and SB. Implementations of the SI and SB factors in this study are limited and not meant to establish new measures, but rather to fulfill a need to generate representative measures based on the collected questionnaire data. By conducting the EFA across several validated questionnaires sensitive to suicide, we were able to establish a more comprehensive understanding of SI and SB. Future work would benefit from using this as empirical support for the mechanistic differences between SI and SB when developing suicidality studies, rather than the direct implementation of these measures.

### Time-Frequency Analyses

Utilizing these SI and SB factors, we conducted TF analyses looking across frequency bands, which is rarely done. Previous studies have looked into time-domain components (Alison et al., 2021; Tsypes et al., 2019; Albanese et al., 2019; Song et al., 2019), individual frequency measures (Tsypes et al., 2021), or ideation and attempts individually (Tsypes et al., 2021; Tsypes et al., 2019), but none, to our knowledge, have looked comprehensively across all frequency measures in the context of both SI and SB.

TF analyses offer the ability to parse out details that could not be seen in time-domain data (Bernat et al., 2015; Watts et al., 2018; Buzzell et al., 2022). By looking at SI and SB separately, we found that across all but one frequency band (alpha), the feedback responses of those with SB was stronger to overall gambling feedback (both losses and gains).

#### Delta and Theta Effects

For delta and theta, we see significant loss and gain responses, and, differently from the HFs, we see significant loss-gain differences.

Increased delta-FN/P3 in the SB group suggests increased orienting to gambling feedback for both gain and loss. Increased theta-FN in those with SB suggest increased salience to loss relative to gain stimuli (Bernat et al., 2015). Delta-P3/SW findings reflect increased evaluative processing of both gain and loss feedback stimuli for those with increased SB, but with significantly more evaluation of loss outcomes than gain. This unique relationship with loss feedback in delta-P3/SW and theta-FN is supported by previous work showing significantly lower feedback-related delta power to gain versus loss in suicide attempters compared to a non-suicide attempters group (Tsypes et al., 2021). Other work done in processing of negative and positive material between healthy controls and those with remitted depression (RMD) found increased P3b and SW in the RMD group for negative material (Liu et al., 2017).

LF regressions (Tables 5 and 6) revealed that delta-FN/P3 effects were accounted for by delta-P3/SW, but theta-FN was a unique predictor of SB. In a relevant study with anxious participants during a gambling task, delta-FN, a component of interest in the gambling task (Ellis et al., 2018), accounted for a majority of the effects. Although we see similar delta and theta effects to that study, the delta-P3/SW measure identified in this study accounts for the delta effects. This could be indicative of prolonged or lengthier processing of the task in those with SB compared to those with SI, or even those with anxiety.

#### Beta-1, Beta-2, and Gamma Effects

For most of the HFs (i.e., beta-1, beta-2, and gamma), SB, but not SI, demonstrated significant relationships to both loss and gain feedback, with no loss-gain differences. Because regression analyses indicated that the individual HF bands did not contain unique information relative to SB, we combined them together into a single HF factor for ease of interpretation.

While the amplitude of the HF factor was increased for both loss and gain, it was increased more for the gain than loss, resulting in significant loss-gain differences for the HF factor that were not seen for the separate frequencies. This was also seen in the individual frequency correlations for loss-gain differences, with negative (though, not significant) correlations indicating a stronger gain effect.

To better understand the HF factors, iot is important to discuss what the activity in each frequency band may indicate. Previous work has indicated that beta activity is a dominant brain wave in people who are alert, anxious, or have their eyes open, and is most frequently picked up fronto-laterally in a symmetrical distribution (Satapathy et al., 2019). This aligns with the current study’s findings of the fronto-temporal and symmetrical beta-1 and beta-2 activity beginning shortly after feedback. Since this effect occurs for both loss and gain trials, it is possibly related to general alertness or increased anxiety across all feedback. There is potential for this to relate to anxiety, as previous work has a found a relationship with suicidality and anxiety (Nepon et al., 2010). Gamma waves were found to be involved in attention, working memory, and long-term memory processing (Malik et al., 2017). In this task, gamma may be related to attention processing or working memory. Reward processing work with beta and gamma suggests that this could be a response to unexpected gains or losses, though effects were localized medial-frontally (HajiHosseini et al., 2012; Marco-Pallarés et al., 2015). The gamma activity could be indicative of a response to any feedback without expectation. Altogether, this may be general response to feedback, or other sensory processes.

#### SB Relationship with All Frequencies

Those with SB may be allocating more attentional and/or decision-making resources toward task feedback compared to those with SI. This differs from other findings indicating that those with SB had deficits in attentional control, memory and working memory compared to depressed individuals and healthy controls (Kelip, Grunebaum, Gorlyn et al., 2012). Work related to decision-making processes offers insight into resource allocation: this may reflect impaired decision-making and processing of subsequent feedback. Previous work suggests that suicide attempters have impaired decision-making based on feedback comparative to healthy or depressed patients (Jollant et al., 2005; Kelip, Gorlyn, Russell et al., 2013; Gorlyn et al., 2013). While other work suggests that impaired decision-making may increase suicidal risk above that conferred by depression (Pustilnik et al., 2017). There is likely a unique effect in those with SB that relates to feedback sensitivity, but more work needs to be done to fully understand why this may be the case, and if similar effects can be seen in other cognitive and affective tasks.

#### Alpha Effects

For alpha, there was a unique response in those with SI towards feedback relative to those with SB. Previous work has localized alpha activity in those with suicidality to the parieto-occipital region (Iznak et al., 2021). They differently indicated that those with suicidal attempts had higher alpha-2 (9-11 Hz) power than those with non-suicidal self-injuries, though both groups presented with similar depression scores. Differently from our study, their recording was conducted in a state of quiet wakefulness with eyes closed. Alpha activity has been shown to reflect a relaxed wakefulness state, decreasing with concentration, stimulation or visual fixation (Stern et al., 2013), alpha may have been suppressed in those with SB compared to those with SI. This could also explain the correlation between alpha and SI indicating some (but not full) suppression of the band. Recent work indicates that relaxation techniques do, to varying degrees, improve anxiety, distress, and depression symptoms indicating a higher need for relaxation in such populations (Hamdani et al., 2022). Since, to our knowledge, no work has been done examining alpha band activity relative to suicidality or depression, research understanding feedback processing would be helpful in understanding these relationships better. Altogether, in the context of this study, this work relating alpha to increased relaxation allows us to infer that the weak alpha relationship with SI suggests high SI individuals as slightly more relaxed relative to high SB individuals.

### Limitations

Although these findings present an exciting direction and new picture of SI and SB, there are limitations. First, the study conducted was correlational, so causal interpretations are not possible. Identifying risk factors is a key gap that new suicide research must address (Acosta et al., 2012; Klonsky & May, 2010, 2014; Naifeh et al., 2019); therefore, it is essential to utilize these results to explore key mechanisms in SI and SB development. Second, this study looked at a singular cognitive task. It would be beneficial to look across other feedback processing and cognitive tasks. Third, although the SI and SB factors are representative of their respective characteristics, they should not be treated as validated measures, but as means to separate suicidality (for the purposes of this study). Despite these limitations, future studies could use these findings as a basis to better understand SB and SI distinctions.

### Conclusions

Overall, the study provides a comprehensive frequency-based outline of the mechanism differences between SI and SB that can be used as a base to conduct future studies and should be validated through other feedback processing tasks. Importantly, this adds to an understood view that it is vital to differentiate between SI and SB to better interpret their significances.

## Supporting information

Supplemental Syntax

## Data Availability

All data produced in the present study are available upon reasonable request to the authors.

## Notes

This work was supported by the United States Department of Defense grant DOD W81 XWH-10-2-0181.

We have no known conflicts of interest to disclose.

### Competing Interest Statement

The authors have declared no competing interest.

### Funding Statement

The study was funded by a Department of Defense grant.

### Author Declarations

Ethics committee/IRB of Florida State University gave ethical approval for this work.

