## Supplemental Syntax for "Feedback Processing as it Relates to Suicidal Ideation and Behavior Using a Time-Frequency Approach"

**Supplementary Materials**

**SPSS Syntax for Suicide Factors**

FACTOR

/VARIABLES BL_BSS_2 BL_BSS_4 BL_BSS_6 BL_BSS_7 BL_BSS_8 BL_BSS_12 BL_BSS_13 BL_BSS_14 BL_BSS_15

BL_BSS_20 BL_BSS_21 BL_SBQR_1 BL_SBQR_2 BL_DSISS_1 BL_DSISS_2 BL_DSISS_3 BL_DSISS_4 BL_SHF_1

BL_SIS_1a BL_BDI2_9

/MISSING LISTWISE

/ANALYSIS BL_BSS_2 BL_BSS_4 BL_BSS_6 BL_BSS_7 BL_BSS_8 BL_BSS_12 BL_BSS_13 BL_BSS_14 BL_BSS_15

BL_BSS_20 BL_BSS_21 BL_SBQR_1 BL_SBQR_2 BL_DSISS_1 BL_DSISS_2 BL_DSISS_3 BL_DSISS_4 BL_SHF_1

BL_SIS_1a BL_BDI2_9

/PRINT INITIAL EXTRACTION ROTATION

/CRITERIA FACTORS(2) ITERATE(25)

/EXTRACTION PAF

/CRITERIA ITERATE(25)

/ROTATION VARIMAX

/SAVE REG(ALL PAFV20_)

/METHOD=CORRELATION.

**SPSS Syntax for Correlations**

NONPAR CORR

/VARIABLES=ideation behavior alphapozloss alphapozgain alphapozlvg beta1loss beta1gain

beta1lvg beta2loss beta2gain beta2lvg gammaloss gammagain gammalvg deltapc1loss deltapc2loss deltapc1gain deltapc2gain deltapc1lvg deltapc2lvg thetaloss thetagain thetalvg

/PRINT=SPEARMAN TWOTAIL NOSIG FULL

/MISSING=PAIRWISE.

**SPSS Syntax for High Frequency Factors**

Loss Trials

FACTOR

/VARIABLES beta1loss beta2loss gammaloss

/MISSING LISTWISE

/ANALYSIS beta1loss beta2loss gammaloss

/PRINT INITIAL EXTRACTION

/CRITERIA FACTORS(1) ITERATE(25)

/EXTRACTION PAF

/ROTATION NOROTATE

/SAVE REG(ALL HFLoss_)

/METHOD=CORRELATION.

Gain Trials

FACTOR

/VARIABLES beta1gain beta2gain gammagain

/MISSING LISTWISE

/ANALYSIS beta1gain beta2gain gammagain

/PRINT INITIAL EXTRACTION

/CRITERIA FACTORS(1) ITERATE(25)

/EXTRACTION PAF

/ROTATION NOROTATE

/SAVE REG(ALL HFGain_)

/METHOD=CORRELATION.

Loss-Gain Trial Differences

FACTOR

/VARIABLES beta1lvg beta2lvg gammalvg

/MISSING LISTWISE

/ANALYSIS beta1lvg beta2lvg gammalvg

/PRINT INITIAL EXTRACTION

/CRITERIA FACTORS(1) ITERATE(25)

/EXTRACTION PAF

/ROTATION NOROTATE

/SAVE REG(ALL HFLvG_)

/METHOD=CORRELATION.

**Stata Syntax for Robust Regressions**

rreg behavior deltapc1loss deltapc2loss

rreg behavior deltapc1gain deltapc2gain

rreg behavior deltapc1lvg deltapc2lvg

rreg behavior deltapc2loss thetaloss

rreg behavior deltapc2gain thetagain

rreg behavior deltapc2lvg thetalvg

rreg behavior deltapc2loss thetaloss hfloss_1

rreg behavior deltapc2gain thetagain hfgain_1

rreg behavior deltapc2lvg thetalvg hflvg_1
